# Translating 3D Slicer into Brazilian Portuguese: A methodological approach to software localization in Latin America

**DOI:** 10.1101/2025.09.15.25335771

**Authors:** Paulo Eduardo de Barros Veiga, Luiz Otavio Murta, Douglas Samuel Gonçalves, Lucas Sanchez Silva, Víctor M. Montaño-Serran, Enrique Hernandez-Laredo, Valeria Gómez-Valdes, Monserrat Ríos-Hernández, Andras Lasso, Steve Pieper, Adriana Vilchis-González, Sonia Pujol

## Abstract

3D Slicer is an open-source software platform for the analysis, segmentation, and three-dimensional visualization of medical imaging data. Although the platform is used by an international research community, its interface was historically available primarily in English, which may limit accessibility for non-English-speaking users. This study describes the development of an ad hoc methodology for the Brazilian Portuguese localization of 3D Slicer within the broader Latin American localization initiative. The methodology addresses recurrent linguistic challenges identified in a preliminary corpus of 300 interface strings, including domain-specific vocabulary, acronyms, word order, passive voice, syntagms, and the adaptation of technical terms. The translation process emphasizes textual uniformity, cohesion, terminological accuracy, and contextual validation in biomedical-computational environments. The proposed framework may support similar software localization efforts in other non-English-speaking contexts, especially when technical precision and linguistic adaptation must be balanced.

## Introduction

The multilingual and multicultural nature of healthcare presents challenges in the usability of medical software. Language barriers prevent comprehension and effective use. Translating and localizing interfaces improves navigation, reduces errors, and promotes broader adoption, considering “the engagement of regional partners in the adaptation and contextualization of the programs” [1]. Providing software in user-preferred languages enhances trust, engagement, and respect for cultural diversity.

In this context, biomedical translation is complex because it depends on nuanced terminology, disciplinary conventions, and interface-specific uses of language. Automated tools may overlook subtleties, cultural nuances, and idiomatic expressions, leading to misunderstandings [2]. Errors in menus, prompts, or interface messages can compromise user interpretation and may increase the risk of misuse in biomedical-computational environments, particularly when machine translation is used without domain-sensitive human review [3, 4]. AI systems and large language models (LLMs) also struggle with domain-specific jargon in medicine [5]. Thus, although LLMs have made remarkable progress in automated translation, their use in critical domains such as medicine requires deliberate methodological frameworks and sustained human intervention through safe translation practices. This approach does not diminish the value of AI, which is useful for translation tasks and large volumes of text; rather, it ensures that these capabilities are applied judiciously, promoting linguistic responsibility—a key concept in this approach.

The use of a foreign language in medical software increases cognitive load, affecting comprehension and decision-making [6, 7]. The Foreign Language Effect demonstrates that second-language use leads to more analytical but less emotional decisions, resulting from distinct neural processing [8]. Neuroimaging confirms that foreign language processing activates different brain areas [9], which impacts tasks such as reading, visual search, and interaction with complex terminology. Language also shapes health-related decisions, such as care-seeking behavior among medical students, influenced by linguistic and grammatical structures. Understanding these effects supports better design for non-native English users.

Concerning this linguistic-computational scenario, this article presents the translation process of the software 3D Slicer into Portuguese, initiated in July 2023, with a focus on an ad hoc methodology designed to ensure textual uniformity, cohesion, and accuracy. By reducing errors and expediting revision, the method offers a systematic approach to software translation, contributing a novel procedural framework.

3D Slicer is a free, open-source, and comprehensive software for analyzing 3D medical image datasets, serving as a hub of a diverse global community. It is intended for use by researchers, the medical community, and developers, facilitating efficient improvement and distribution of new methods to clinical users. However, 3D Slicer is not authorized for clinical use and is intended for research purposes. The software enables easy package installation through its Python and C++ extensibility, and serves as a Jupyter kernel with remote 3D rendering capabilities. 3D Slicer enables analysis, segmentation, and visualization of medical images, demonstrating versatility for various types of organ data [10, 11]. Despite its versatility, language barriers may hinder its accessibility, limiting its adoption among non-English-speaking researchers and healthcare professionals.

The Brazilian Portuguese localization was developed within the broader Slicer for Latin America initiative, which provides public resources for Portuguese and Spanish localization, tutorial development, documentation, workshops, and community training [12]. The initiative includes translation files, glossaries, translated tutorials, documentation, workshop materials, and repositories supporting the adoption of 3D Slicer by Latin American research and training communities [13, 14].

In this context, translating medical software like 3D Slicer poses linguistic challenges due to its technical nature and multilingual context. Key issues include domain-specific vocabulary, acronyms, word order, passive voice, syntagms, and the adaptation of foreign terms. Each demands targeted solutions to preserve clarity, precision, and cultural sensitivity, especially given that 3D Slicer is not merely a tool, but a multidisciplinary, intercultural community.

Domain-specific terms must avoid ambiguity; acronyms require contextual adaptation; and structural shifts—such as adjusting passive constructions and syntagms—ensure fluency in Portuguese. Balancing accuracy and cultural-linguistic coherence is essential when integrating foreign terminology.

Addressing these demands involved both linguistic and technical expertise, supported by collaboration with experienced colleagues and specialized tools. These elements were key to developing the ad hoc methodology presented here.

The following sections will outline the materials and methods used to establish this methodology, detailing the linguistic resources employed, the solutions developed, and the structured approach taken to ensure translation accuracy.

## Materials and methods

A specific methodology was developed to translate 3D Slicer into Brazilian Portuguese within the broader Slicer for Latin America initiative. This methodology represents an ad hoc process characterized by its object-oriented and problem-sensitive approach. The proposed linguistic topics may vary depending on the translation context and the nature of the software being translated. Therefore, it is not an a priori methodology, but a method built through engagement with the linguistic object and with the technical constraints of the software interface.

The Brazilian Portuguese translation workflow covered several 3D Slicer components and associated resources, including the core interface, CTK, LanguagePacks, MONAILabel, SlicerIGT, SlicerVMTK, TutorialMaker, and the project glossary. The translation and linguistic revision process was carried out by the Brazilian team translator in coordination with technical contributors. Translation decisions were supported by terminology research, glossary consultation, contextual verification in the 3D Slicer interface, and comparison with related documentation and tutorials.

The preliminary corpus of 300 English interface strings was assembled during the ongoing localization workflow. It was not designed as a random or statistically representative sample, but as an exploratory and problem-oriented corpus. Strings were selected when they appeared grammatically, terminologically, or contextually relevant to the development of a translation methodology. The initial corpus was drawn from 5,382 3D Slicer source strings available at that stage of the project; it did not include Slicer CTK, LanguagePacks, MONAILabel, SlicerVMTK, or SlicerIGT, which were incorporated into subsequent localization stages. The 300-string subset (5.57% of the source strings considered at that stage) was recorded in an internal spreadsheet together with the adopted Portuguese translation and a brief linguistic comment. These comments identified issues such as passive voice, acronym treatment, word order, syntagmatic organization, domain vocabulary, or lexical adaptation. This procedure made it possible to identify recurrent patterns and formulate the flowchart-based translation guidelines presented in this article. The translation process therefore involved dictionaries, grammar resources, academic databases, specialized literature, and contextual comparison within the software environment. These procedures established guidelines for the translation effort, helped reduce inconsistencies during revision, and contributed to the construction of a reproducible localization method [15].

One key component in the translation of medical software, including 3D Slicer, is the implementation of a comprehensive glossary. A glossary is vital to medical software systems because it promotes standardized communication, improves user experience, supports training, contributes to data accuracy, facilitates compliance, and favors interoperability [16]. In this project, the glossary included recurrent terms central to 3D Slicer and medical image computing, such as *bounds, coordinate system, fiducial, IJK, labelmap, MRML, RAS, segmentation, transform, volume*, and *voxel* [14]. Medical terminology is complex and filled with jargon; therefore, a glossary helps users, researchers, developers, and healthcare professionals share a consistent understanding of the terms used within the system. It also reduces the learning curve for new users and offers a stable reference for unfamiliar terms. Since medical terminology and practices evolve, the glossary can be updated as part of the software translation process, remaining integrated into the methodological framework presented in this article.

Contextual validation was an essential part of the workflow. The translator used the 3D Slicer interface and contextual string-search resources to locate translated or untranslated expressions in their actual interface environment. This step helped determine whether the proposed Portuguese translation was appropriate for menus, buttons, prompts, tutorials, and other interface elements. Technical and domain-specific issues were discussed with members of the biomedical and computational teams when necessary, particularly for terms related to medical imaging, segmentation, label maps, voxels, spatial orientation, and computational procedures.

Regarding the Portuguese language, the methodology identified six key linguistic topics relevant to software translation and outlined primary challenges based on the sampled data. As detailed below, a flowchart was developed for each topic, constituting a central component of the methodology. The diagrams were designed to enhance the visualization of the translation process by applying basic principles of algorithmization to systematize the sequence of stages. The process begins with an example-problem or problem-sentence, representing the challenge the translator must tackle. This methodology was developed to solve textual problems while standardizing the quality of translations in a biomedical context. From the problem-sentence, a translation hypothesis is formulated—either more literal or contextually driven—enabling a flexible translation approach. This hypothesis is then tested against the broader textual and technical context of the field, including specialized literature, dictionaries, glossaries, repositories, and actual software usage. Every translation hypothesis must be evaluated in relation to this broader network of texts and interface contexts, where the accuracy and usability of the translation can be assessed. The diagrams guide the process by questioning the occurrence of similar translations within the relevant field, thus arriving at an occurrence-based solution for the problem-sentence. This approach aims to ensure high-quality and contextually safe translations, particularly in the sensitive domain of biomedical texts. [17]

The sampling effectively identified key translation issues and facilitated the formulation of solutions to optimize the process. These topics encompass domain vocabulary, acronyms, word order, passive voice, syntagms, and Portuguese linguistic adaptation. Other issues, such as verbal omission, could be included. However, because of their linguistic obviousness, no processes or patterns are developed for these topics, even though they are part of the translation task. One may also note that in software translation, it is essential to consider not only the verbal elements but also the visual layout of the 3D Slicer interface. Nonverbal aspects often play a key role in shaping meaning and guiding user interaction, binding the pragmatic and semantic levels. After all, meaning is also influenced by the spatial arrangement and interactive design in which language and interactions appear.

### Linguistic tools

The comprehensive analysis of 300 translated sentences aimed to identify patterns between various linguistic elements, including specific domain terminology, acronyms, sentence structure, verb forms, adaptation of foreign words, syntactic nuances, and phrase organization. Linguistic tools have facilitated the standardization process and solved linguistic issues. Therefore, it is frequently included in flowcharts.

Throughout the translation process, cross-referencing of the text ensured lexical accuracy and accommodated newly introduced terms or neologisms. The *Vocabulário Ortográfico da Língua Portuguesa* (VOLP) [18] was used as the authoritative lexical reference for Portuguese. In addition, dictionaries such as the Houaiss dictionary [19], the Oxford Dictionary [20], and the public 3D Slicer glossary [14] were consulted. To validate translations, an extensive search was conducted in academic repositories [21–23], among others, reviewing articles and theses from Brazilian and international universities, focusing on academic texts in the relevant field. This methodology ensured linguistic precision and facilitated comparative analyses with other languages, notably Spanish and French.

Additionally, contextual string-search resources [24] were instrumental in identifying specific phrases within the project context. These resources allowed for the location and comparison of sentences within the software environment, providing a valuable means of verifying translations. In pursuit of improved accuracy, contextual checking proved essential for achieving semantic precision and maintaining alignment with the software’s visual design. These materials form the essential linguistic tools used when the flowcharts refer to “linguistic tools.”

## Results and flowcharts

To address the challenges of translating open software such as 3D Slicer, a structured methodology was developed, applying principles of algorithmization to optimize the translation process, as illustrated in the flowcharts presented in this paper. This methodology frames each linguistic issue as a “problem-sentence”, formulates translation hypotheses, and iteratively tests these hypotheses within broader contexts. Integrating linguistic tools, algorithmic approaches, and collaborative efforts—bringing together Mexican and Brazilian teams in an international setting—ensures accurate and contextually appropriate translations. Beyond improving quality and consistency, this methodology may enhance usability, facilitate adoption, and reduce risks associated with linguistic ambiguity in biomedical-computational applications.

Despite structural and cultural differences between Portuguese and Spanish, the ability of Portuguese-speaking team members to understand written and spoken Spanish significantly facilitated communication and comprehension. This linguistic affinity not only streamlined the exchange of information but also made possible a sense of cultural cohesion within the Latin American teams. In this collaborative environment, the didactic contributions of the Mexican team proved particularly valuable, enabling the Brazilian team to navigate challenges more effectively. As a result, this integrative translation approach strengthened cross-cultural connections within the 3D Slicer environment, ultimately enhancing users’ linguistic experience.

Specifically, this section outlines the established patterns for each linguistic topic identified through the ad hoc methodology. It presents the linguistic topics identified through the sampling process, accompanied by synthetic textual explanations and schematic figures, without aiming to create an exhaustive grammar list. The topics are domain or field vocabulary, acronyms, word order, passive voice, syntagms, and Portuguese linguistic adaptation. Each flow chart describes the translation processes. This was the most concise way to present the methodology.

### Domain or field vocabulary

Domain vocabulary refers to specialized and unique terms for specific fields or professions. Translators, professionals, and scholars in specialized areas, particularly those concerning the 3D Slicer and its array of biomedical-technological terms, must accurately employ the appropriate vocabulary. Misapplication of these terms can lead to misunderstandings and compromise the integrity of the translated content.

Considering this vocabulary issue, the scheme depicted in Fig. 1 focuses primarily on ensuring domain-specific translation within a particular area of expertise. This approach ensures the precise treatment of technical and specialized vocabulary during translation, thereby preventing terminological distortions and upholding the conceptual fidelity, nuances, and standards specific to the field. This is an essential pattern when translating software.

**Fig 1.**
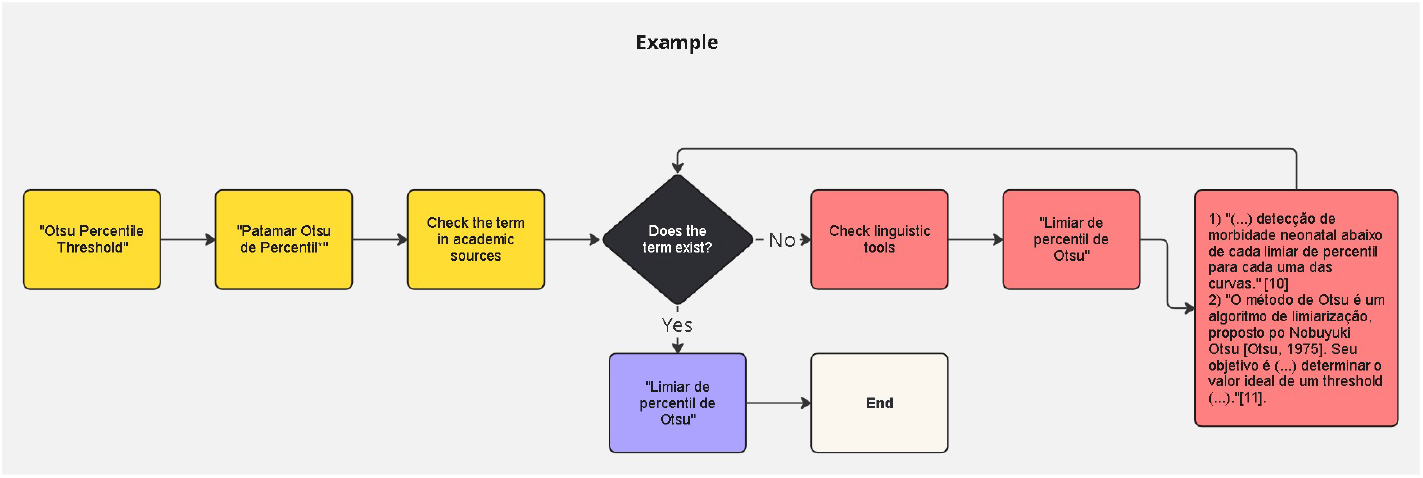
Pattern—Domain vocabulary. Flowchart for translating domain-specific technical terms. The process begins with a problem-sentence, proposes a translation hypothesis, verifies field-specific usage in academic or technical sources, and revises the solution with linguistic tools when necessary.

The diagram in Fig. 1 outlines a translation process that begins with the problem-sentence the linguist will face: “Otsu Percentile Threshold”, a technical term. The first step is to propose a linguistically more flexible translation, forming the basis for the hypothesis: “Patamar Otsu de Percentil” in Portuguese. This hypothesis is then verified in academic sources to determine whether it appears in the broader textual and technical context of the field. If the term is found *ipsis litteris*, it is confirmed, and the process ends. Linguistic tools are consulted if the term is not found or is found only partially, resulting in a revised translation. Therefore, “Limiar de percentil de Otsu” is supported by references to academic texts that examine the concept and Otsu’s threshold algorithm. For additional examples, 3D Slicer contains numerous technical terms from the biomedical field, creating an exhaustive list that would be impractical to detail.

In most cases, the literal or direct translation aligns with the terminology commonly used in the field. For instance, “Feret diameter in mm” was translated as “Diâmetro de Feret em milímetros”. In such cases, the translator had to cross-reference the term within its specific context, using the step-by-step diagram as a methodological guide.

### Acronyms

Translating acronyms poses a challenge and holds significant importance in ensuring accurate comprehension in translations. [25] While more common in English, using acronyms is less prevalent or even avoided in Portuguese. Acronyms represent linguistic constructs specific to a particular language and culture, often deeply embedded in social and professional contexts. Merely substituting them in the target language, altering only the order of letters, can result in loss of meaning or misinterpretation. Therefore, the precise translation of acronyms is paramount for preserving terminological accuracy and cultural connotations, thereby safeguarding the translation’s integrity and communicative effectiveness, regardless of the sentence’s length or the display of buttons in 3D Slicer.

The two flowcharts in Fig. 2 explain the methodological process of translating acronyms from English to Brazilian Portuguese. The first flowchart outlines the general steps: locate the acronym, translate it literally or nearly so, verify if the acronym exists or functions in Portuguese, and then decide whether to keep it in English, use it in Portuguese, or fully translate it into a non-acronym form. The second flowchart provides a specific example drawn from the 3D Slicer context, illustrating the application of this process. It demonstrates how to address the challenges of handling acronyms in languages that culturally use them in very different ways, depending on whether they are used in Brazilian Portuguese or should remain in English due to field-specific conventions or bibliographic references. There are numerous examples of acronyms throughout 3D Slicer. The use of acronyms is indeed significant and culturally impactful for Portuguese speakers. Beyond the example described in the diagram, we also encounter the occurrence of “FA” in a biomedical imaging context, which represents Fractional Anisotropy. Translating it into Portuguese as “AF” (Anisotropia Fracionada) would create unnecessary confusion since the medical field is more accustomed to “FA” in English than to “AF.” The best solution to cater to a broad 3D Slicer audience is to resolve the acronym by spelling out “Anisotropia Fracionada.” Similar cases occur with “DTI” (Diffusion Tensor Imaging), translated as “Imagem por Tensor de Difusão,” and “PET” (Positron Emission Tomography), translated as “Tomografia por Emissão de Pósitrons.” When text length reduction is needed, keeping the acronym in English is preferable.

**Fig 2.**
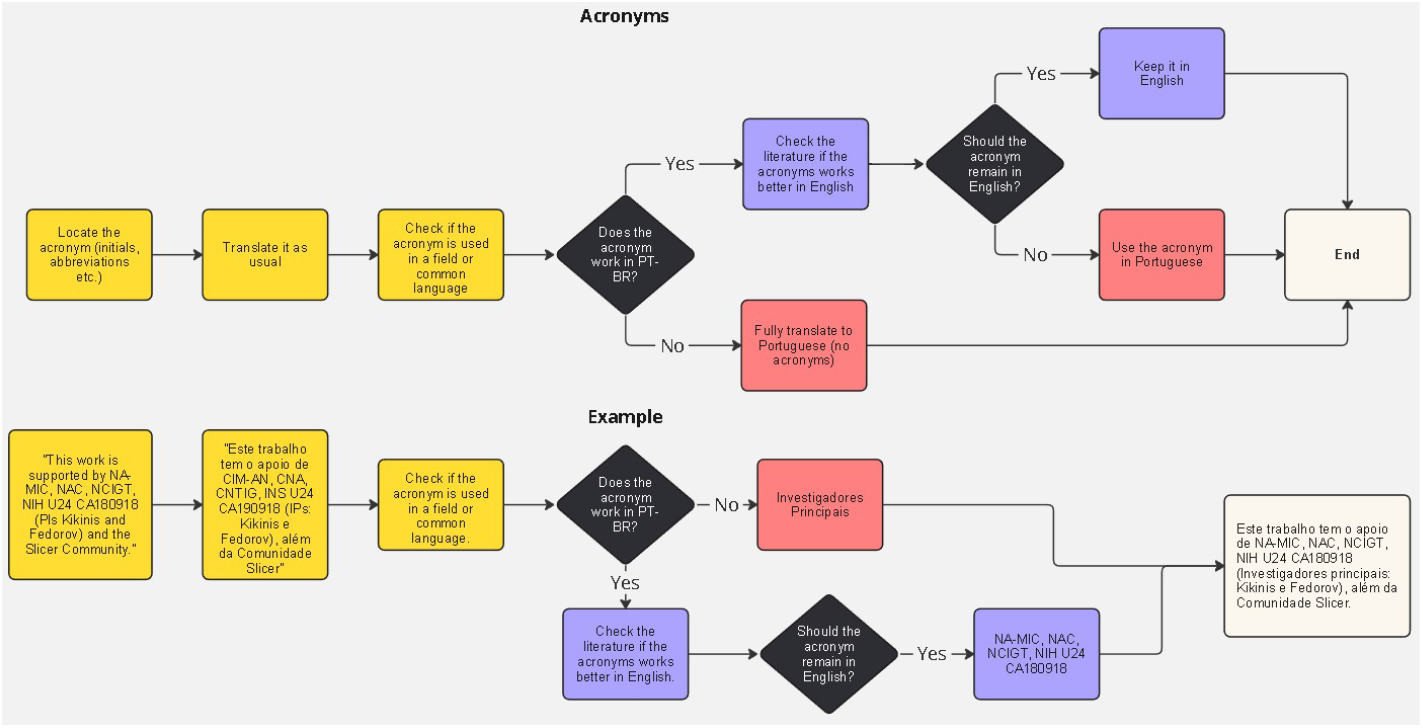
Pattern—Acronyms. Flowcharts for deciding whether an English acronym should be retained, translated as an acronym, or expanded in Brazilian Portuguese. The process balances field-specific convention, clarity, and target-language usage.

### Word ordering

Considering word order in translation is crucial due to structural differences between languages, which can impact clarity and coherence. [26] In this context, word ordering refers to the determination of the most appropriate syntactic sequence during translation. English typically follows the adjective-noun order, while Portuguese favors the noun-adjective order, resulting in reversed ordering rules, as a pattern that can also be observed. Simply substituting words may not be enough to maintain the original meaning; reordering is often necessary for comprehensibility and naturalness. Sometimes, a significant structural change in the sentence may be needed. Neglecting word order can lead to awkward or incoherent translations. Adapting sentence structure according to target language conventions ensures effective communication.

Fig. 3 provides a structured approach to ensuring that translations maintain syntagmatic coherence and semantic integrity when transitioning into Portuguese. Furthermore, according to the diagram, the notation “(((Closing) (%1)) ((running) (modules)))” is demarcated with brackets to delineate syntagmatic blocks or segments, reflecting a simplified syntagmatic structuring model discussed in the linguistic literature [27]. This segmentation reveals the following syntagmatic levels: (Closing), (%1), (running), (modules) at the base level; (Closing %1) and (running modules) at an intermediate level; and finally, (Closing %1 running modules) as the fully integrated structure.

**Fig 3.**
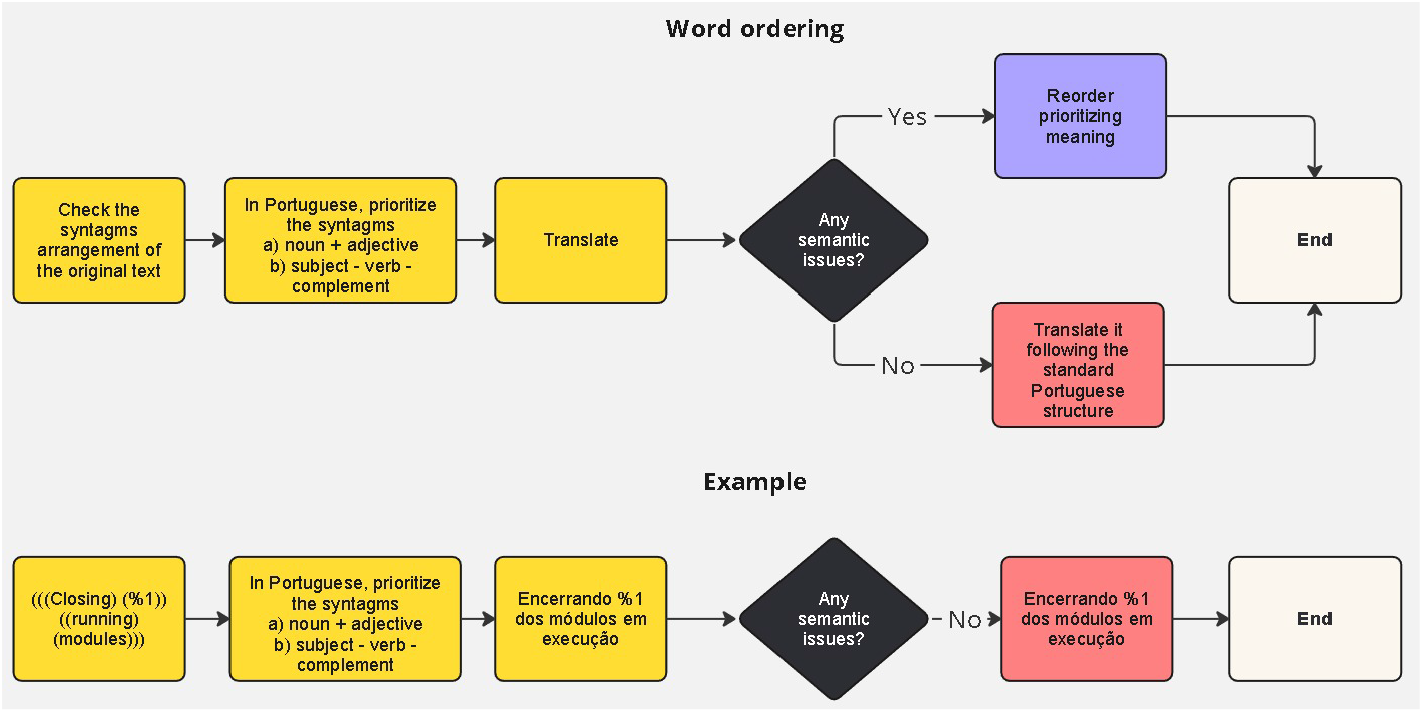
Pattern—Word ordering. Flowchart for reorganizing English syntagmatic sequences according to Portuguese word-order conventions while preserving semantic clarity in interface strings.

Another example is the original sentence: “Confirm source representation change.” The best approach is to preserve the syntagmatic ordering of Portuguese during translation, yielding: “Confirmar a alteração da representação da fonte.” In Portuguese, adjectives often need to be transformed into adjectival phrases, using a preposition followed by a noun, to make the sentence more natural to native speakers. All methods presuppose such adjustments to ensure fluency and a natural flow for the native ear. Translation is always an exercise in good judgment, one that goes beyond rigid rules. Generally, in terms of sentence structure, Portuguese tends to adopt the reverse order of English, with the syntagmatic order being translated inversely. For instance, “harden regularization transform” is most naturally translated as “transformação de regularização robusta.” In this case, the first word in English becomes the last in Portuguese, and so forth.

Additionally, it is crucial to follow nominal regency, which often requires the inclusion of a preposition and, when necessary, a definite article in Portuguese. However, this is not always the case. Some sentences maintain the same order in translation, such as “Make source” which is translated as “Gerar fonte” where the first word in English corresponds directly to the first word in the Portuguese translation, as well as the last one. There are also hybrid sentences where part of the syntagmatic sequence is preserved, including the first word. In contrast, another part is reordered, as in “Update %1 representation using custom conversion parameters” translated as “Atualizar a representação %1 utilizando parâmetros de conversão personalizados”. The issue of sentence ordering must be considered in all translated sentences, except for those involving single words or untranslated elements such as command lines (“SlicerCapture.avi”; “SlicerCaptureLightbox.png”; “H.264” etc.).

### Passive voice

The use of passive voice in English and Portuguese exhibits variations primarily in their respective frequencies of use, rather than in structural disparities. In both languages, the passive voice is constructed using the form of the verb “to be” followed by the past participle of the main verb. However, while the passive voice is more prevalent in English, particularly in formal contexts, Portuguese tends to favor the active voice to express similar notions. [28] Furthermore, Portuguese grammar allows one to use the “synthetic passive voice,” which is more prevalent and notably more concise in such contexts. Consequently, linguistic usage emerges as a determinant in this context, warranting careful consideration according to the mother tongue, thus underscoring its significance as an essential linguistic pattern.

The flowchart in Fig. 4 outlines a decision-making process for translating English sentences in the passive voice into Portuguese. Initially, the flowchart branches depending on whether the English sentence is identified as being in the passive voice. If the sentence is in the passive voice, the translation begins with an assessment to determine if a synthetic passive construction is more appropriate in Portuguese. Given the grammatical flexibility of Portuguese in this topic, both synthetic (using the pronoun “se”) and analytic passive constructions are valid options. If the synthetic passive is suitable, the sentence is adjusted accordingly; otherwise, the analytic passive is maintained. On the other hand, if no passive voice is detected in the English sentence, a direct translation in the active voice is performed.

**Fig 4.**
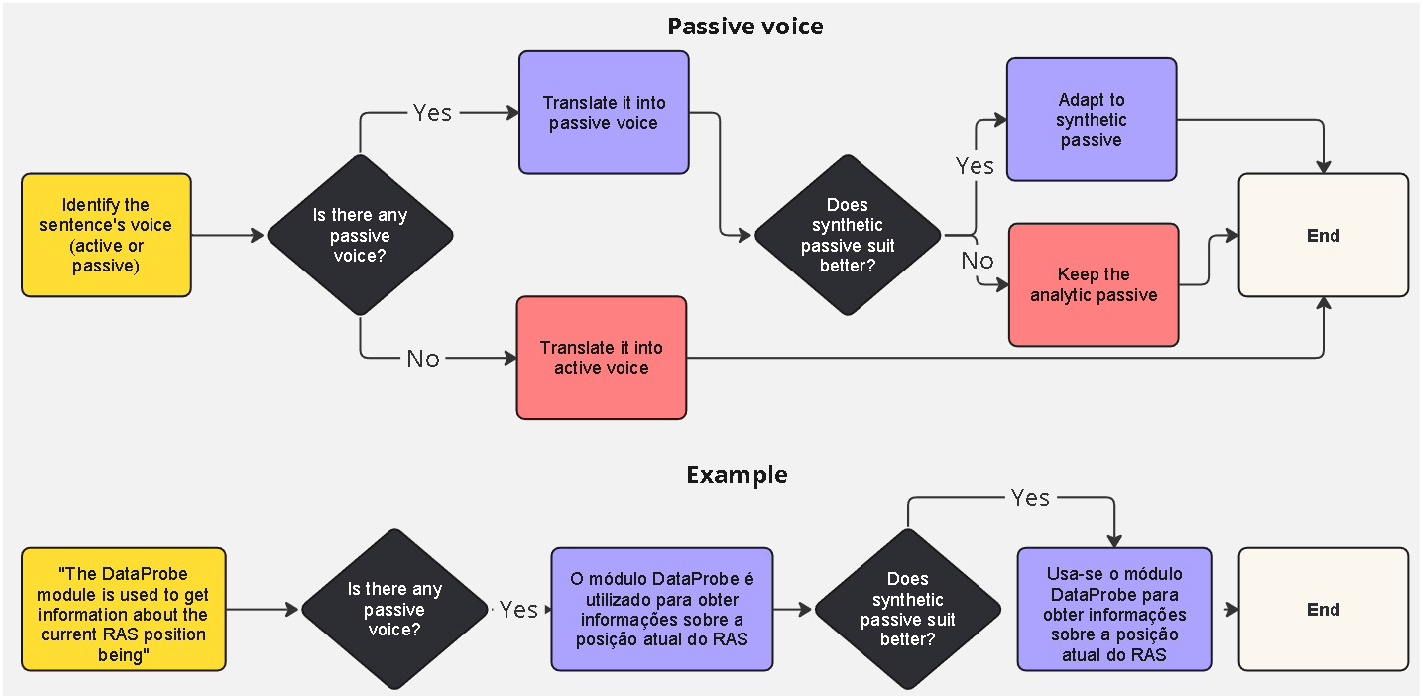
Pattern—Passive voice. Flowchart for translating English passive constructions into Portuguese, including the choice between analytic passive, synthetic passive with *se*, and active-voice reformulation.

For example, the English sentence “The DataProbe module is used to get information about the current RAS position” can be translated into Portuguese using the analytic passive and then adapted to a synthetic passive structure. This adaptation often results in a more natural and concise translation, reflecting the grammatical options in Portuguese and challenging the common belief that English is inherently more synthetic than Portuguese. The final Portuguese rendering, “Usa-se o módulo DataProbe para obter informações sobre a posição atual do RAS” exemplifies this transformation. It is worth noting that while the synthetic passive is sometimes debated as a form of active voice, this analysis adheres to the traditional normative perspective, given ongoing discussions in grammar. Another example is the sentence: “Location, size, and shape of initial segments and content of source volume are taken into account.” In this case, the synthetic passive is the most appropriate choice for a smooth and concise translation. The Portuguese equivalent, “Consideram-se a localização, o tamanho e a forma dos segmentos iniciais e o conteúdo do volume de origem” highlights the importance of verb agreement with plural subjects in Portuguese. In cases where an entirely passive meaning is required in the target language and the synthetic passive is not suitable, the analytic passive takes precedence. For instance, the sentence “This work was funded by CCO ACRU and OCAIRO grants” is translated into Portuguese as “Este trabalho foi financiado pelos subsídios de ACRU, CCO e OCAIRO” preserving the passive voice.

### Syntagms

In generative grammar, a syntagma refers to a syntactic unit comprising one or more words that function together as a cohesive unit. It typically includes a headword or phrase that dictates the nature of the syntagma, accompanied by elements such as nouns, adjectives, verbs, adverbs, and prepositions. Analyzing syntactic units can aid in translation tasks by enhancing comprehension of sentence structure, maintaining cohesion and coherence, selecting appropriate equivalents, and facilitating cultural adaptation.

The image in Fig. 5 presents a flowchart that outlines the translation of syntagms from English to Portuguese. It begins by analyzing the syntagmatic structure in English, with a particular focus on specific concepts within the relevant field. Next, the complete translation of the syntagms is attempted based on the English sentence, using linguistic tools to scan for syntagmatic patterns. If an exact match for the full syntagm exists, the translation follows the standard terminology used in the field of knowledge. However, if the full syntagm does not exist, the process involves breaking the expression into smaller syntagms, checking each one individually, and rearranging them until the correct terms are identified. Once the syntagms are located, they are reassembled, completing the translation. For example, after analyzing its syntagmatic components, the phrase “Standard deviation of scalar values” is translated as “Desvio padrão de valores escalares.” If needed, the syntagms are separated and then correctly recombined during the process.

**Fig 5.**
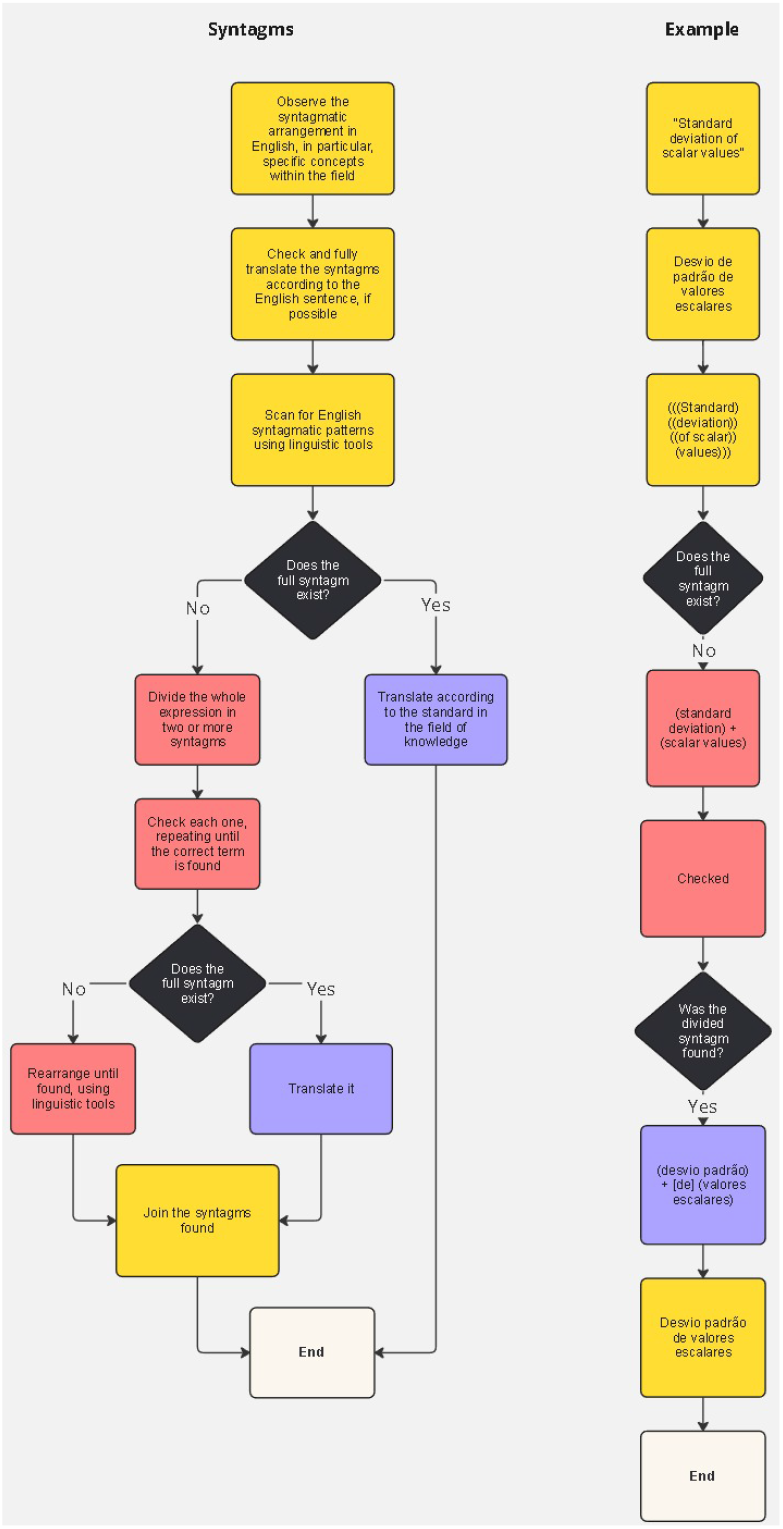
Pattern—Syntagms. Flowchart for analyzing technical syntagms, identifying standard equivalents in the field, and recombining translated components into coherent Portuguese expressions.

**Fig 6.**
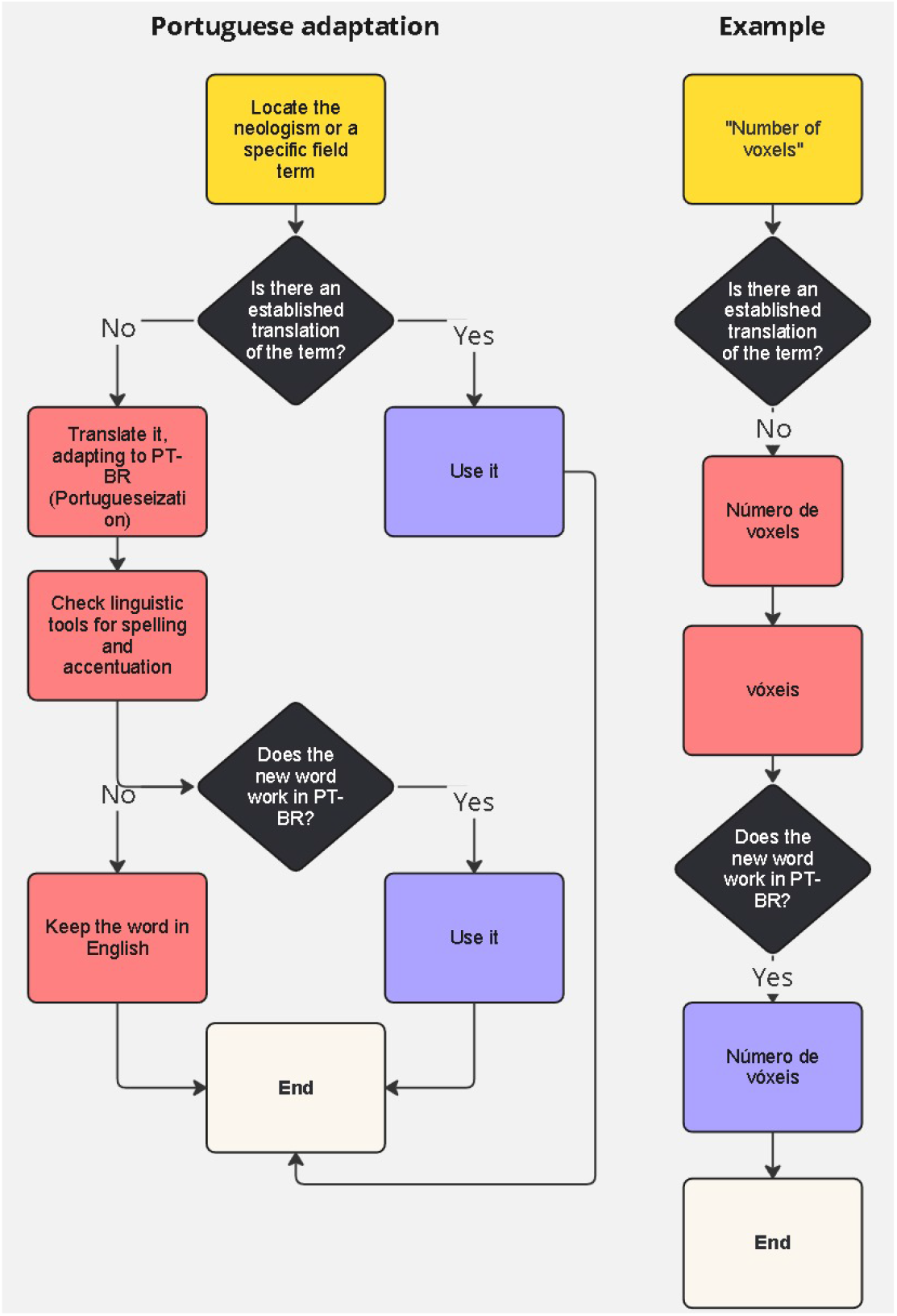
Pattern—Portuguese adaptation. Flowchart for adapting neologisms and specialized English terms into Brazilian Portuguese when established equivalents are unavailable or insufficient.

The critical issue in the diagram in Fig. 5, which should not be confused with sentence ordering, is finding the exact terms in the biomedical-computational literature. Extended expressions, during translation, can often become confusing in the resulting text. Breaking down the syntagms to find the correct corresponding terms is necessary, ensuring they align with the specific area of expertise. Another example is the sentence “Change source representation to binary labelmap?” where the syntagms need to be grouped to identify pairs of terms that function together and should remain connected. Notably, “source representation” (representação de origem) and “binary labelmap” (mapa de rótulos binário) must be kept as grouped pairs for an accurate translation.

### Portuguese adaptation

Adapting foreign terms to one’s native language, with a preference for the latter, is fundamental for effective communication and the preservation of linguistic identity. It is imperative to thoroughly prioritize Portuguese terms as a guiding principle. However, when established Portuguese equivalents are unavailable or would distort specialized usage, retaining foreign-language terms may be necessary to avoid artificial or misleading adaptations. This approach fosters clarity and ensures that language reflects the cultural nuances of its speakers. This proposal aligns with the objectives of “3D Slicer for Latin America: Localization and Outreach,” emphasizing the importance of linguistic authenticity and cultural preservation.

The image illustrates a flowchart for adapting neologisms or specialized field terms from English to Brazilian Portuguese (PT-BR). The process begins by identifying the neologism or specific term. If an established translation exists, it is used directly. If not, the term undergoes translation and adaptation into PT-BR. Next, linguistic tools are consulted to verify the correct spelling and accentuation. If the adapted word fits PT-BR standards, it is used. If it does not, the original English term is retained. For example, “Number of voxels” is first translated as “Número de vóxeis.” Since the adapted word works within PT-BR linguistic conventions, it is used to complete the process. The same principle applies to the word “pixels,” which becomes an adapted term in Portuguese through accentuation, resulting in “píxeis.” A notable example is the sentence “Set reference image geometry and resamples all segment label maps,” where the English term “resample” is used. In informal Portuguese, the neologism “reamostra” and its variants (reamostragem, reamostrar) could be used. However, the Vocabulário Ortográfico da Língua Portuguesa (VOLP), a key tool for confirming officially registered words, omits “reamostrar.” Although the rule is to primarily follow the VOLP, in this case, due to the contingent use of the term, we adopt the neological use of “reamostrar” and its variants.

### Possible flowcharts

While other topics, like verbal omission, could have been considered, their exclusion is justified by their inherent linguistic clarity. Although not addressed, the absence of a formal process or pattern for these topics does not detract from the overall translation task. The development of a flowchart illustrating the system of verbal omission would reveal that, for specific grammatical issues, the flowchart would be so concise that a predefined pattern would be unnecessary. In such cases, the absence of a formal pattern would ensure quality and consistency in translation. Flowcharts were only created for linguistic circumstances where standardization would enhance the rigor of the translation, ensuring uniformity and textual quality. For example, compared to the other charts, only a basic flowchart on verbal omission is provided as an example of its dispensability.

Verbal omission, a common linguistic phenomenon in both English and Portuguese, involves the exclusion of certain verbs from sentences without compromising their grammatical integrity or clarity of meaning. In Portuguese, the omission of verbs can occur more frequently and across various sentence types, reducing sentence length and complexity. This linguistic feature enables Portuguese speakers to convey information more concisely and efficiently, facilitating smoother communication and enhancing comprehension of written and spoken discourse.

Translators must consistently consider the following step: ‘Can any verbs be omitted?’ This is especially pertinent for state verbs and gerunds. Such consideration can lead to shorter sentences in Portuguese than in English without compromising clarity or fidelity to the original text.

The flowchart in Fig. 7 outlines a decision-making process for determining whether verbs can be omitted in a given translation task. It begins by instructing the user to identify all verbs and gerunds in the source text. The next step presents a decision point: if the meaning of the text can be conveyed without using any verbal or verbal-nominal forms, the verbs can be omitted. However, if removing the verbs would compromise the meaning or grammatical structure of the sentence, all verbs must be retained. This flowchart provides a clear framework for translators to consider when evaluating verb omission, a complex aspect of translation. State verbs such as “ser” and “estar” (to be, in English) can often be omitted to avoid unnecessarily lengthening sentences, especially in software translation. Another example: “Video creation failed: ffmpeg executable path is invalid: *{*path *}*.” In this case, omitting the verb (is) makes the sentence more direct and concise: “Falha na criação do vídeo: caminho executável ffmpeg inválido: *{*path*}*.”

**Fig 7.**
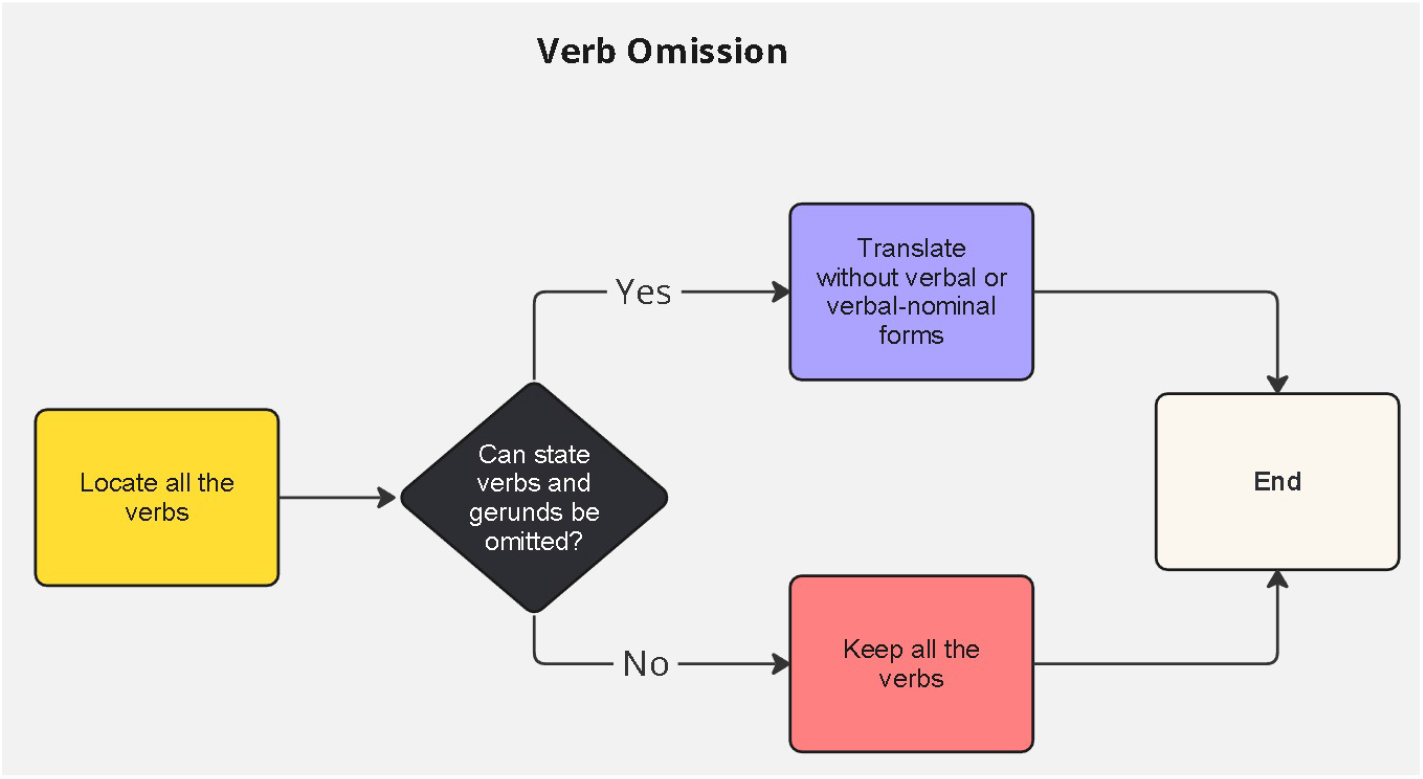
Pattern—Verbal omission. Flowchart for deciding whether state verbs and gerunds can be omitted in Portuguese without loss of meaning, especially in concise interface strings.

## Discussion

This study presents the Brazilian Portuguese localization of the open-source medical image processing software 3D Slicer and, more specifically, the methodological framework developed to address recurrent linguistic problems in the translation process. The proposed method was built from the practical demands of the localization workflow and organized around six main topics: domain-specific vocabulary, acronyms, word order, passive voice, syntagms, and Portuguese linguistic adaptation. Verbal omission was also considered as a secondary issue, although it did not require the same degree of formalization. The work is connected to public localization resources, including translation files, glossaries, documentation, tutorials, and workshop materials developed within the Slicer for Latin America initiative [12–14].

The main contribution of the study is not the claim that a single universal method can solve all problems of software translation. Rather, the contribution lies in the systematization of a case-sensitive procedure in which interface strings are treated as linguistic problems requiring contextual validation. This procedure combines terminological research, comparison with academic and technical sources, glossary construction, and evaluation of the interface context. In this sense, the method is ad hoc, but not arbitrary: it is built from the object being translated and from the recurrence of identifiable linguistic patterns.

Compared with more literal approaches to software translation, the proposed method prioritizes accuracy, fluency, and terminological stability. Literal translation may be useful in many technical contexts, especially when established terminology already exists in the target language. However, literal equivalence is insufficient when English syntactic ordering, acronyms, neologisms, or passive constructions produce unnatural or ambiguous Portuguese interface strings. The methodology therefore requires the translator to test each translation hypothesis against specialized usage, linguistic tools, and the pragmatic conditions of the software interface.

The treatment of acronyms illustrates this point. Some acronyms should be preserved in English because this is the form recognized by the biomedical field, while others may be translated or expanded to avoid ambiguity for Portuguese-speaking users. Similarly, syntagmatic analysis and word-order adjustment are necessary because English and Portuguese organize technical noun phrases differently. These issues are not peripheral; in software localization, short interface strings may contain dense technical information, and small linguistic distortions can affect comprehension.

The approach also has practical implications for collaborative localization projects. By creating recurrent patterns and flowcharts, the method supports consistency across translators and reviewers and reduces the number of isolated decisions made during revision. This is especially relevant in multilingual and international projects such as 3D Slicer for Latin America, where Brazilian, Mexican, and North American teams must coordinate linguistic, biomedical, and computational expertise.

The study has limitations. The initial analysis was based on a subset of 300 interface strings, corresponding to 5.57% of the source strings considered at that stage. This corpus was exploratory and problem-oriented rather than random or statistically representative. Although it was sufficient to identify recurrent translation problems and to develop initial patterns, additional modules and later translation stages may present further linguistic issues. The methodology should therefore be understood as expandable and revisable rather than final. Future work may test the same framework across other 3D Slicer modules, compare Portuguese and Spanish localization patterns, and evaluate user comprehension before and after localization.

Finally, the implications of this work should be understood within the research-oriented status of 3D Slicer. The Portuguese localization may improve accessibility, training, adoption, and usability for researchers and professionals working in biomedical imaging contexts. However, it should not be interpreted as a direct clinical intervention or as evidence of improved diagnostic outcomes. Its primary contribution is linguistic, methodological, and infrastructural: it helps make a research software environment more accessible to Portuguese-speaking users while preserving technical precision.

## Conclusion

The translation of 3D Slicer into Brazilian Portuguese contributes to the accessibility of open-source biomedical software in Latin America and demonstrates the importance of linguistic methodology in software localization. The project addressed not only language barriers, but also the need for cultural and terminological adaptation in a complex biomedical-computational environment.

The ad hoc methodology developed in this study proved useful for organizing translation decisions, ensuring textual uniformity, improving cohesion, and reducing inconsistencies during the revision stage. The analysis of 300 interface strings allowed the identification of six central translation patterns: domain vocabulary, acronyms, word order, passive voice, syntagms, and Portuguese linguistic adaptation. These patterns provided a practical framework for guiding the translation of similar strings throughout the localization process.

The collaboration among Brazilian, Mexican, Canadian, and American teams was essential to the development of the project. This international setting made it possible to combine linguistic, biomedical, and computational expertise, while also strengthening the Latin American dimension of the localization initiative. The integration of linguistic tools, glossary construction, contextual validation, and flowchart-based decision patterns supported a more systematic approach to software translation.

The study shows that software localization in biomedical contexts requires more than lexical substitution. It demands attention to syntax, terminology, interface context, disciplinary usage, and the target language’s own expressive resources. The methodology presented here may therefore serve as a reference for future localization projects involving research software, especially when technical accuracy and linguistic naturalness must be balanced.

By making 3D Slicer more accessible to Portuguese-speaking users, this work contributes to broader participation in biomedical imaging research and training. It also reinforces the need for collaborative and methodologically explicit approaches to the translation of scientific software.

## Data Availability

All data produced are available online at www.slicer.org

## Data availability statement

The data underlying this study consist of 3D Slicer interface strings, translation examples, localization resources, and glossary entries used in the Brazilian Portuguese localization workflow. Public localization files and related resources are available through the 3D Slicer and Slicer for Latin America repositories and project pages [12–14]. The 300-string exploratory corpus used for methodological analysis was maintained as an internal working spreadsheet during the translation process; representative examples from this corpus are included in the manuscript and figures. No patient-level, clinical, or personally identifiable data were used in this study.

## Acknowledgments

We sincerely thank Elena Nascimento Veiga for her valuable assistance in reviewing the text and engaging in linguistic discussions related to English translation and academic writing.

